# Micro-RNAs miR-375-3p and miR-7-5p are released alongside ACTH from corticotroph pituitary neuroendocrine tumor

**DOI:** 10.1101/2021.06.17.21259073

**Authors:** Helvijs Niedra, Raitis Peculis, Ilze Konrade, Inga Balcere, Mihails Romanovs, Liva Steina, Janis Stukens, Jelizaveta Sokolovska, Janis Klovins, Vita Rovite

**Author notes:** **Corresponding author**: Vita Rovite.

## Abstract

**Objective:** Circulating miRNAs are found in bodily fluids including plasma and can serve as biomarkers for diseases. The aim of this study was to provide the first insight into the landscape of circulating miRNAs in close proximity to the adrenocorticotropic hormone (ACTH) secreting PitNET. To achieve this objective next-generation sequencing of miRNAs in plasma from bilateral inferior petrosal sinus sampling (BIPSS) - a gold standard in diagnosing ACTH-secreting PitNETs, was carried out.

**Methods:** Sinistral (left) and dextral (right) BIPSS blood samples of the patient were collected in three time points: before the administration of corticotropin-releasing hormone, 5 and 15 minutes after stimulation. Peripheral venous blood samples were also collected 24 hours before and after BIPSS and before the resection of PitNET and 24 hours after. In differential expression analysis sinistral plasma was compared with dextral.

**Results:** BIPSS concluded that the highest amount of ACTH was released in the sinistral side at the 5th minute mark indicating a presence of tumor. The highest amount of differentially expressed miRNAs was observed 5 minutes after stimulation (20 upregulated, 14 downregulated). At the 5^th^ minute mark in sinistral plasma, two miRNAs were identified: hsa-miR-7-5p and hsa-miR-375-3p that were highly upregulated compared to other BIPSS samples and peripheral plasma samples. Clustering analysis showed that BIPSS plasma differs from peripheral plasma in miRNA expression patterns.

**Conclusions:** data indicates that ACTH-secreting PitNET actively releases two circulating miRNAs upon stimulation with CRH (hsa-mir-7-5p, hsa-mir-375-3p) alongside with ACTH implying further studies of these miRNA as diagnostic markers are needed.

## 1. Introduction

ACTH-secreting PitNETs are responsible for the majority of ACTH-dependent Cushing’s syndrome (CS) cases (75 - 80%) while the other 15% - 20% can be classified as CS with an ectopic origin (Sharma et al., 2015). It is also estimated that only 6% - 9% of ACTH-dependent CS cases can be attributed to large (>10 mm diameter) ACTH-secreting tumors (Woo et al., 2005), (Katznelson et al., 1998). To prescribe appropriate treatment it is vital to correctly assess the main cause of ACTH-dependent CS and as first step patients are required to undergo magnetic resonance imaging of the pituitary gland (MRI) (Newell-Price et al., 2006). However, the study by Invitti et al. which investigated PitNETs cases that are less than 5 mm in diameter, it was concluded that only 53% of MRI scans yield conclusive results (Invitti et al., 1999). In a CS scenario when the results of MRI scan are inconclusive in confirming the presence and location of a tumor the patients may be required to undergo bilateral inferior petrosal sinus sampling (BIPSS) procedure [4]. BIPSS is a gold-standard procedure for distinguishing between pituitary and non-pituitary sources of ACTH-hypersecretion. It involves simultaneous catheterization of inferior petrosal sinuses (IPSs) which directly drain the pituitary gland. The catheterization itself is done via the transjugular approach and after the position of both catheters is adjusted a simultaneous administration of desmopressin or corticotropin-releasing hormone (CRH) is performed. This stimulates the corticotrophs of the pituitary gland and PitNET to release ACTH. After the stimulation of corticotrophs, blood samples are taken directly from IPS and concentrations of ACTH are measured. Depending on the setting this diagnostic approach has up to 100% sensitivity and specificity in identifying whether the source of ACTH hypersecretion is the PitNET and on which side (sinistral or dextral) the tumor is located (Jarial et al., 2018; Zampetti et al., 2016).

miRNAs are a class of small non-coding RNAs with an average length of 22 nucleotides. miRNAs play a vital role in post-transcriptional gene expression regulation by binding to the 3’ UTR region of target mRNAs and up to 60% of human protein-coding genes contain conserved targets for miRNA-mRNA interaction (Arteaga-Vázquez et al., 2006; Friedman et al., 2009; Wightman et al., 1993). For this reason they, have been extensively studied in the context of cancer development, progression, diagnostics, and therapy with a vast amount of miRNAs showing promising results as both tissue biopsy and liquid biopsy biomarkers (Shigeyasu et al., 2017; Tan et al., 2018). Regarding PitNETs miRNAs also have been studied both in tumor tissue and as blood-based biomarkers. In tissue, it has been shown that upregulation of miR-34a and miR-107 is directly associated with downregulation of AIP expression, one of the most studied genes in familial isolated PitNETs and a proposed tumor-suppressor (Beckers et al., 2013; Dénes et al., 2015; Trivellin et al., 2012). In ACTH-secreting PitNETs miR-26a has been found to be significantly upregulated compared to normal pituitary tissues with its direct target being cell cycle regulation associated protein kinase PRKCD (Gentilin et al., 2013).

As of today, there is a limited amount of information on circulating extracellular vesicle-associated miRNAs in blood plasma of PitNET patients. Nemeth K. *et al*. were the first ones to provide insight into potential plasma-derived miRNA biomarkers for PitNET with miR-143-3p emerging as a candidate biomarker for post-surgery monitoring of FSH/LH PitNET patients (Németh et al., 2019). Another study showed that plasma miRNAs: miR-16-5p, miR-145-5p and let-7g-5p are upregulated in CS patients with ACTH-secreting PitNETs versus CS patients with ectopic cause (Belaya et al., 2020).

Detecting PitNET specific miRNAs in plasma samples remains a challenging task due to their high dilution. In this study we had a unique opportunity to study PitNET-derived circulating extracellular vesicle (EVs) associated miRNAs in close proximity to the tumor itself alongside the EVs bound miRNAs in plasma from peripheral venous blood (PVB). We hypothesized that there would be detectable differences in expression of circulating EVs bound miRNAs between sinistral and dextral sides of IPSs as well as differences between BIPSS and PVB plasma. To confirm this hypothesis, we collected plasma samples from the BIPSS procedure in three separate time points and compared the miRNA fractions found in PVB using NGS analysis.

## 2. Materials and methods

### 2.1. Patient recruitment and blood sample processing

The patient was recruited from Riga East Clinical University Hospital where she underwent BIPSS in 2018. Before the acquisition of biological samples and clinical data two written informed consents were obtained from the patient: consent for the use and storage of biological material in the national biobank Genome Database of Latvian population (LGDB) for health and hereditary research and PitNET research specific consent (Rovite et al., 2018). The use of patient’s samples and data in this study was approved by the Central Medical Ethics Committee of Latvia (protocol: 22.03.07/A7 and 2/18-02-21).

BIPSS was done by an experienced team of interventional radiologists and endocrinologists using a percutaneous femoral vein approach. After obtaining basal samples, an intravenous injection of 100mcg human CRH (hCRH) was administered as a bolus, and post-stimulation samples were obtained at 0, 3, 5, 10, and 15 minutes. All samples were immediately collected in pre-chilled tubes containing EDTA and kept on ice until centrifugation. During the BIPSS procedure, the blood samples for miRNA extractions were also collected in EDTA tubes in three main steps (Supplementary Figure 1). Two months after BIPSS patient underwent transsphenoidal surgery during which the medical staff collected PVB samples before surgery and 24 hours after surgery. All samples used for miRNA isolation were stored at RT until centrifugation. The plasma was separated from whole blood directly after sample collection by two-step centrifugation: 2000 RPM for 10 minutes at RT and 4000 RPM for 10 minutes at RT. After centrifugation, the plasma samples were aliquoted at 1ml and stored at −80 °C.

Plasma located EVs were isolated and their RNA contents were extracted from 0.5 – 1ml plasma samples using exoRNeasy Midi kit (Qiagen, Germany) according to manufacturer’s instructions. During the extraction 52 synthetic miRNA spike-ins from QIAseq miRNA Library QC qPCR Assay Kit (Qiagen, Germany) were added according to the manufacturer’s instructions before the RNA purification part. Extracted RNA was divided into two aliquots – one for NGS library preparation and one for quality control analysis by qPCR. Aliquots were stored under −80 °C.

### 2.2. miRNA library preparation and NGS

To determine the validity of extracted RNA samples prior to downstream NGS analysis we carried out qPCR on ViiA™ 7 (Applied Biosystems, USA) using QIAseq miRNA Library QC qPCR Assay Kit. qPCR reactions were set up following the manufacturer’s instructions. QIAseq miRNA Library QC qPCR Assay Kit included 8 assays which allowed us to assess the following conditions: extraction efficiency and uniformity (UniSp100 and UniSp101), cDNA synthesis efficiency (UniSp6), evaluation of hemolysis (miR-23a-3p and miR-451a), presence of constitutively expressed plasma miRNAs (miR-103a-3p, miR-191-5p, miR-30c-5p).

Samples that passed quality standards underwent miRNA library preparation using Small RNA-Seq Library Prep Kit (Lexogen, Austria) according to the manufacturer’s instructions. To avoid the contamination of adapter dimers and RNAs that exceed the range of small ncRNAs we employed BluePippin electrophoresis (Sage Science, USA) for the size selection step. According to Lexogen’s user manual fragments around 140 – 150 bp indicate a small ncRNA library so the target range for BluePippin electrophoresis was set 125 – 160bp as it was found to be the most optimal for yielding purified library. The results after BluePippin were visualized using High Sensitivity DNA Chip and Kit on Agilent 2100 bioanalyzer (Agilent Technologies, USA) and concentrations were measured using Qubit 2.0 (Thermo Fisher, USA). Libraries were sequenced on the MiSeq NGS system (Illumina, USA) with a target range of 4 million reads per sample. During extraction, library preparation, and sequencing the samples were processed in two batches (Supplementary Table 1).

### 2.3. NGS data analysis and differential expression analysis

The NGS data were analyzed using CLC genomics workbench (v20.0.4.) (Qiagen, Germany). Adapters were trimmed from the 3’ end and any reads with the quality score < 0.02 were discarded. Regarding the length based trimming any reads that did not fall into the range of 15 - 55 nucleotides were discarded. For miRNA and spike-in quantification, the reads to were aligned miRBase (v22) and qiaseq_mirna (v1) spike-ins dataset. For alignment, maximum allowed mismatches were set to 2 and for length-based isomiRs maximum upstream/downstream bases were set to 2. Allowed sequence length was set from 18 to 25 bp (size of mature miRNA). Differential expression analysis was performed in CLC Genomics Workbench (v20.0.4). The methodology is based on negative binomial distribution and uses trimmed mean of m-values (TMM) as normalization method. Count values are output as counts per million (CPM).

For BIPSS samples differential expression analysis we compared the dextral side with the sinistral side in three separate time points: 0 minutes (before administration of CRH), 5 minutes after stimulation, and 15 minutes after stimulation. For PVB samples we compared before BIPSS with after BIPSS and before surgery with after surgery. We chose to represent results that met the following criteria: Bonferroni P < 0,05. Fold changes were represented as Log2 fold change (Log2FC). To validate our results in different sample sets we used publicly available data by Nemeth K. *et al*. In this study the authors also sequenced circulating EV plasma miRNAs obtained from 20 plasma samples. The plasma samples were collected from 10 gonadotropins secreting (FSH/LH) PitNET patients (Németh et al., 2019). Since the available dataset had reads marked with unique molecular indices (UMIs) which are supported by CLC Genomics Workbench (V20.0.4) during miRNA quantification we used UMIs to quantify the miRNAs.

## 3. Results

### 3.1. Patient clinical characteristics

The enrolled patient (female) was 24 years old when the first symptoms of an irregular menstrual cycle, hirsutism, and weight gain developed. The first hormonal measurements were done in late 2015 (Supplementary Table 2.1.) which showed elevated levels of ACTH (66.3 pg/ml, reference range 0 – 46 pg/ml). As the symptoms progressed in 2017 patient was seen by endocrinologist and overnight 1 mg *Dexamethasone* test was done, where no suppression had been reached with cortisol levels of 546 nmol/L. Following this high dose two-day *Dexamethasone* was advised (supplementary table 2.2.). In 2018 the patient underwent dynamic gadolinium-enhanced MRI of the sellar region (Supplementary Figure 2) which demonstrated left-sided microadenoma (0.7 × 0.9 cm in diameter) at the anterior pituitary. Following this laboratory studies were repeated, as a *Dexamethasone* suppression tests (Supplementary table 2.3.). According to the previous laboratory, radiologic and clinical findings, there was an ambiguity concerning low ACTH levels. Consequently, a multidisciplinary team of endocrinologists and radiologists decided to perform BIPSS, to substantiate the source of ACTH as pituitary or ectopic. Elevated central/peripheral ACTH ratio was achieved both before and after hCRH stimulation from the sinistral side, as a result, CS of PitNET origin was confirmed (Table 1).

**Table 1.**
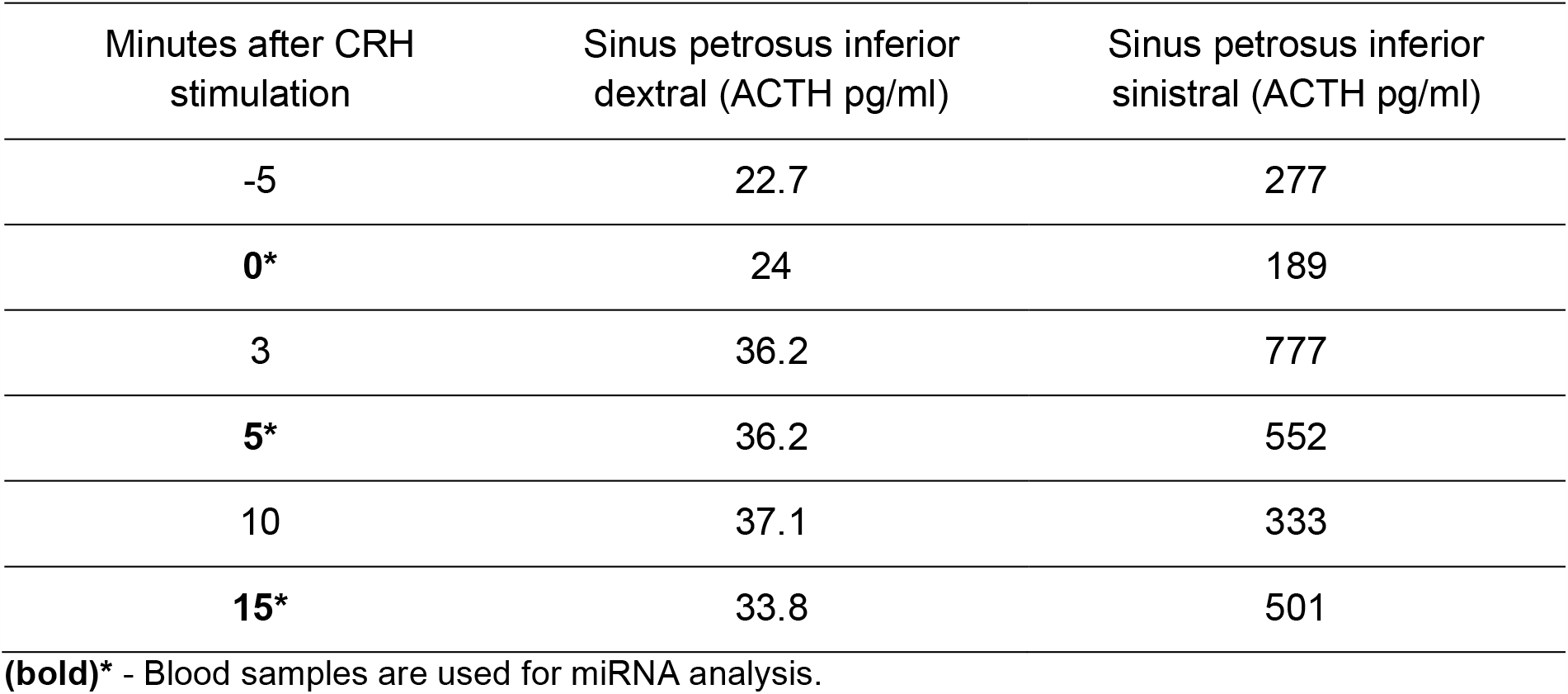
Bilateral inferior petrosal sinus sampling results.

| Minutes after CRH<br>stimulation | Sinus petrosus inferior<br>dextral (ACTH pg/ml) | Sinus petrosus inferior<br>sinistral (ACTH pg/ml) |
| --- | --- | --- |
| -5 | 22.7 | 277 |
| **0\*** | 24 | 189 |
| 3 | 36.2 | 777 |
| **5\*** | 36.2 | 552 |
| 10 | 37.1 | 333 |
| **15\*** | 33.8 | 501 |

**(bold)\*** - Blood samples are used for miRNA analysis.

Transsphenoidal surgery was indicated, and the patient was seen by a neurosurgeon. Consequently, in October 2018, transsphenoidal surgery was successfully performed and the patient was discharged with recommendations to take Hydrocortisone 15 mg divided in two doses.

### 3.2. Quality control of plasma samples and NGS data

For all 10 plasma samples, the Ct value of UniSp6 spike-in was < 20 indicating that no PCR inhibitors were present during QC qPCR (Supplementary Figure 4). For all samples, the ΔCt of miR-23a-3p – miR-451 (hemolysis associated markers) was below 7 confirming that all samples were eligible for further analysis by NGS (Supplementary Figure 5) The samples during RNA extraction were processed in two separate batches. For this reason, a difference in Ct values of spike-ins (UniSp100/101) between some samples was expected to be greater than 3 (Supplementary Figure 3, Supplementary Table 3). Constitutively expressed plasma miRNAs (miR-103a-3p, miR-191-5p, miR-30c-5p) were detected in all samples with Ct values ranging from 24.89 to 36.20. The lowest Ct values were observed in PVB samples (Supplementary Figure 6). On average the sequencing runs yielded 4’009’431 reads per sample (range: 3’468’590 - 5’120’624). Trimming reduced the average read count to 2’830’560 reads per sample (range: 2’151’420 - 3’666’966) (Supplementary Figure 7).

Annotation to miRBase (release v22) for BIPSS samples on average had the annotated reads percentage of - 2.30% (60’884 reads), (range: 1.85 - 3.16%, 41’058 - 95’297 reads). For PVB samples the average annotated read count was 10.98% (336’517 reads) (9.36 - 15.33%, 202’297 - 445’487 reads) (Supplementary Table 4). The difference in annotated read percentage between BIPSS and PVB samples was statistically significant (Welch t-test P = 0,008) As for the quantification of 52 QIAseq miRNA library QC spike-ins there was a noticeable difference between BIPSS and PVB samples before and after BIPSS. For BIPSS samples the spike-in read percentage ranged from 0.16 to 0.40% and for PVB samples this percentage ranged from 1.51% to 2.47%. There was a statistically significant positive correlation between the percentage of spike-in reads and the percentage of annotated miRNAs (Spearman’s rho = 0.75; P = 0.018). According to the spike-ins kit user manual, each sample should correlate with the remaining samples with an R2 of >0.95 (Supplementary Table 9).

### 3.3. Most expressed miRNAs in plasma from BIPSS and PVB

The most expressed miRNA across all six samples from BIPSS was hsa-miR-486-5p which represented 38.3% - 78.7% of annotated counts (average: 62.52%) of all miRNA reads (Figure 1). It was also the most expressed miRNA in plasma samples from PVB: 34.88% in before and after BIPSS samples and 26,95% in before and after surgery samples. There were two miRNAs that were amongst the top 10 most expressed miRNAs in the BIPSS sample group but not in PVB: hsa-miR-10a-5p with an average of 1.06% of total counts and hsa-miR-375-3p with an average of 1.53% of total counts. hsa-miR-375-3p had a considerably small amount of reads (range: 10 - 73) in both BIPSS and PVB except for BIPSS-5-SIN (4581 reads; 9.19% of total counts). Despite having a low percentage of total counts in other BIPSS samples it was still included in the list of 10 most expressed miRNAs within the group of BIPSS samples. Also, for the sample BIPSS-5-SIN, the top 10 most expressed miRNAs had a significantly lower overall percentage (71%) of total aligned counts compared to the other nine samples. This can be attributed to the fact that compared to other BIPSS samples it had the highest amount of unique miRNA species detected (162) (Figure 1).

**Figure 1.**
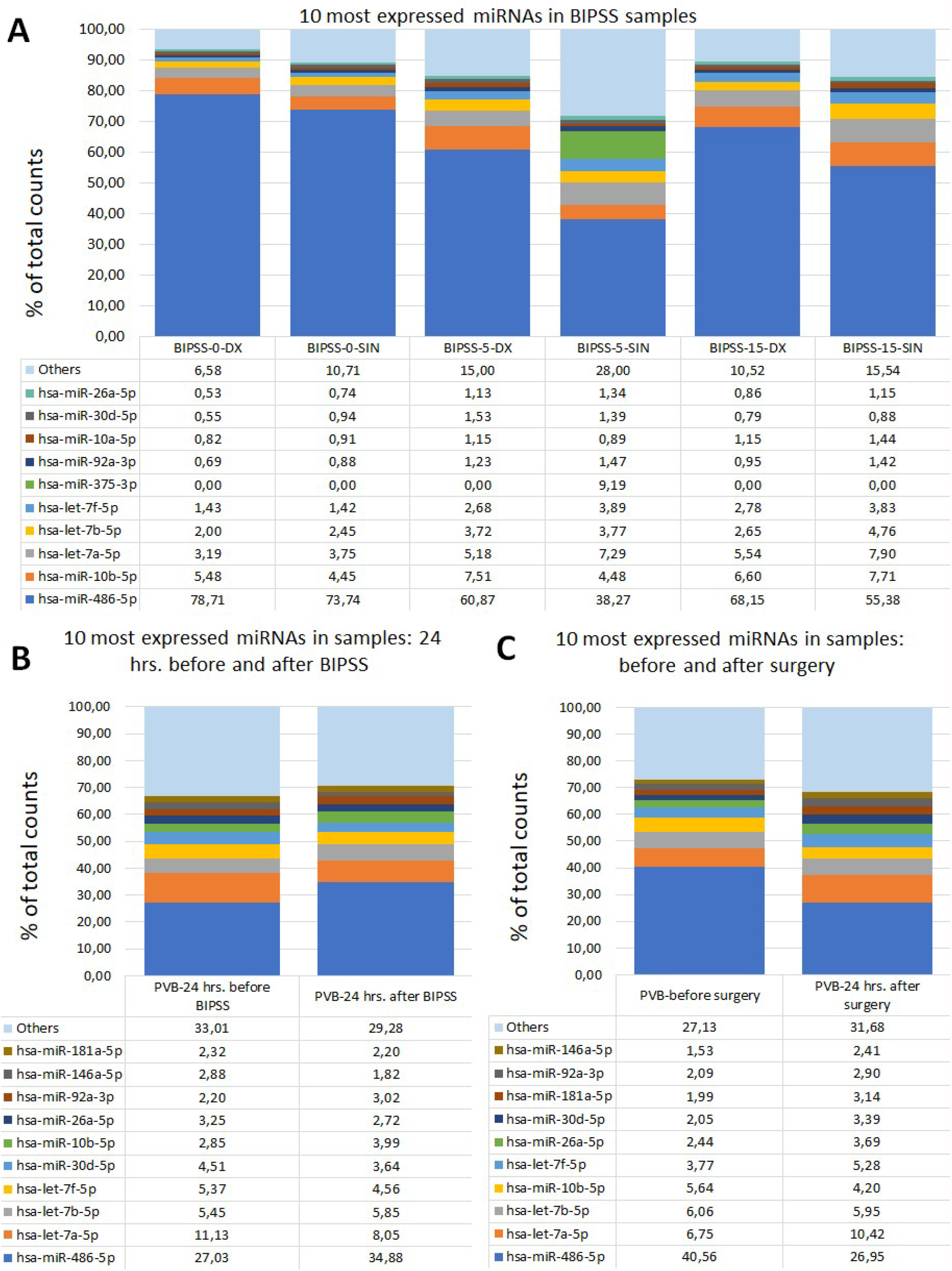
Comparison of the top 10 most expressed miRNAs in BIPSS samples and PVC samples. (A), (B) and (C) graphs represent the most 10 expressed miRNAs and their percentage of total annotated miRNA counts.

### 3.4. Read mapping to human genome

In parallel, we also mapped reads to the human genome (hg38) (Supplementary Figure 8) to evaluate what other types of non-coding RNAs were present within the samples. On average, 85.43% (range: 82.19 - 87.81%) of reads were mapped to hg38 while 14.57% (range: 12.19 - 17.81%) of reads were left unmapped. In the BIPPS sample group on average 23.12% (range: 19.53 - 28.79%) reads were annotated as miRNAs while in the PVB sample group this amount was 28.56% (range: 24.36 - 31.05%). It is interesting to note that BIPSS samples had a higher amount of annotated lncRNA reads 25.21% (range: 18.16 - 31.53%) vs 5.66% (range: 2.98 - 7.10%). PVB samples had markedly higher amounts of miscRNA reads 46.18% (35.42 - 57.89%)

### 3.5. miRNA expression profiles in BIPSS and PVB plasma

We compared miRNA expression patterns of all 10 plasma samples and identified 50 miRNAs whose expression profiles were able to distinguish PVB plasma vs plasma samples from BIPSS (Figure 2). In the clustering analysis, two clusters of samples can be observed: 1st one which represents the plasma from BIPSS containing five of six BIPSS samples, and 2nd one which represents the plasma acquired from PVB containing all 4 PVB samples. The expression profile of the 0 min dextral sample was closer to the 15 min dextral sample than to the 5 min sample. Interestingly, the expression profile of 5 the min sinistral sample was markedly different from all other BIPSS samples including its counterpart BIPSS-5-DX as its expression profile was more closely related to the PVB samples.

**Figure 2.**
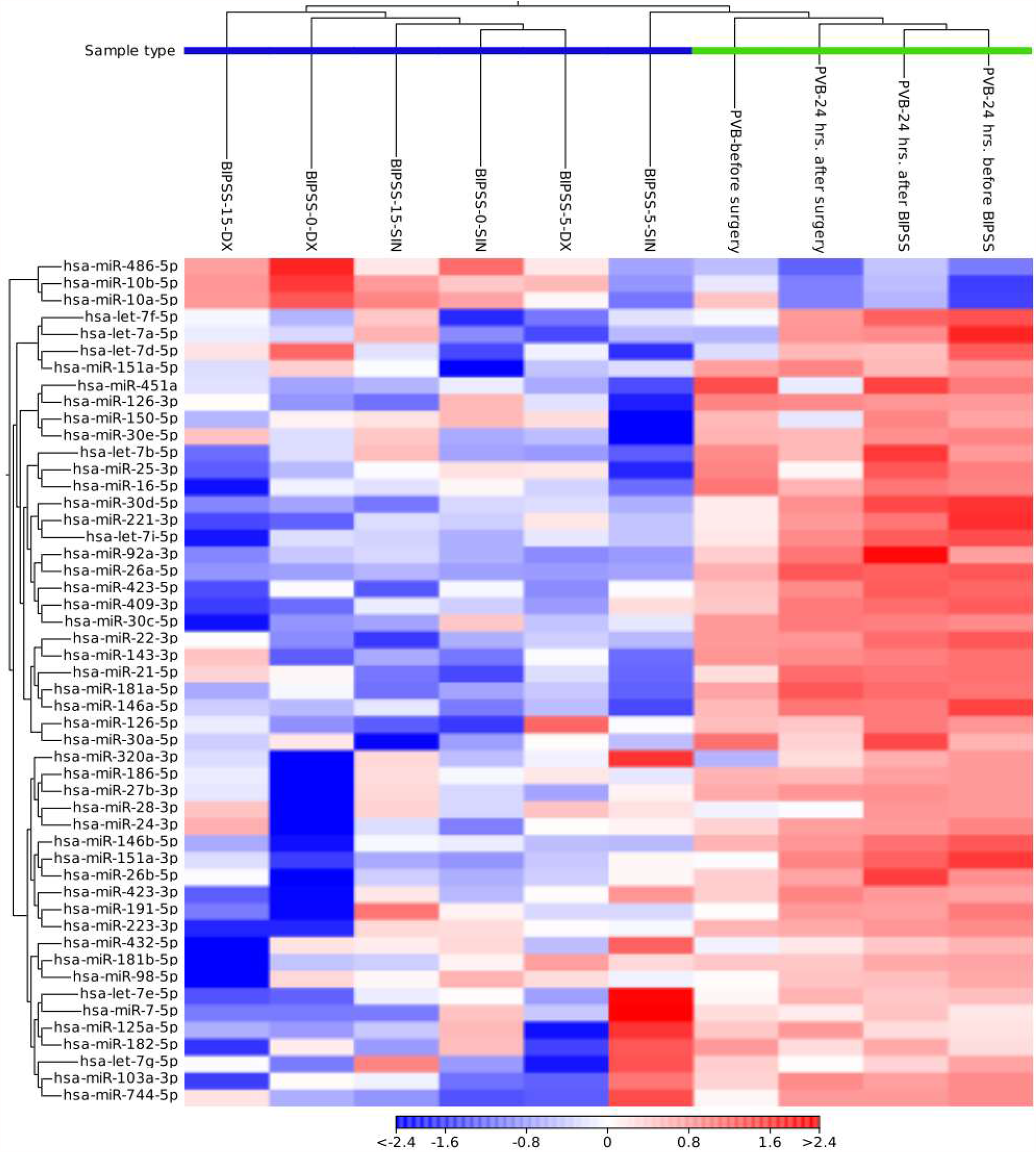
Heat map containing expression patterns of 50 miRNAs to differentiate plasma collected from BIPSS vs PVC. The heat map was generated by selecting a fixed number of features (50) with the highest coefficient of variation and with a minimum supporting count of 10 reads. Distance measure - Euclidean distance. Cluster analysis showed that there is a clear difference in the expression of selected 50 miRNAs between BIPSS and PVC samples. BIPSS-5-SIN sample showed a markedly different pattern than other BIPSS samples.

### 3.6. Differentially expressed miRNAs between sinistral vs dextral side

Analyzing all three time points of BIPSS we identified 49 unique differentially expressed miRNAs (DEMs) of which 12 were repeatedly found in two time points and 32 in only one time point. Five DEMs that were repeatedly differentially expressed across all three time points were: hsa-miR-7e-5p, hsa-miR-423-3p, hsa-miR-486-5p, hsa-miR-409-3p, hsa-miR-126-3p. By examining all three time points together a dynamic can be seen which shows that the highest number of DEMs is present during the 5th minute of BIPSS. The highest Log2FC values were also present within the 5th minute and they ranged from −3.2 to 9.8. In comparison before administration of CRH (0 min), the Log2FC values ranged from −3.8 to 2.7 and during the 15th minute after CRH administration the values ranged from −2.3 to 2.9 (Figure 4), (For more detailed information see Supplementary Table 11).

**Figure 3.**
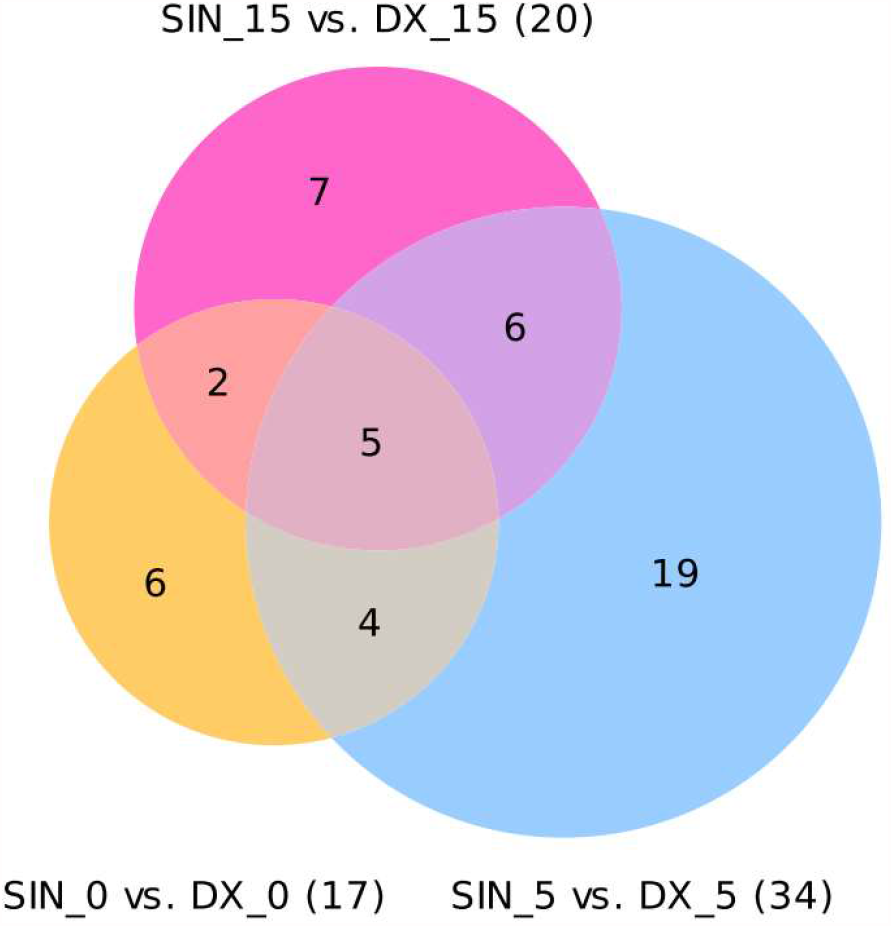
Venn diagram of differentially expressed miRNAs in three time points. The Venn diagram represents differentially expressed miRNAs with a Bonferroni P < 0.05.

**Figure 4.**
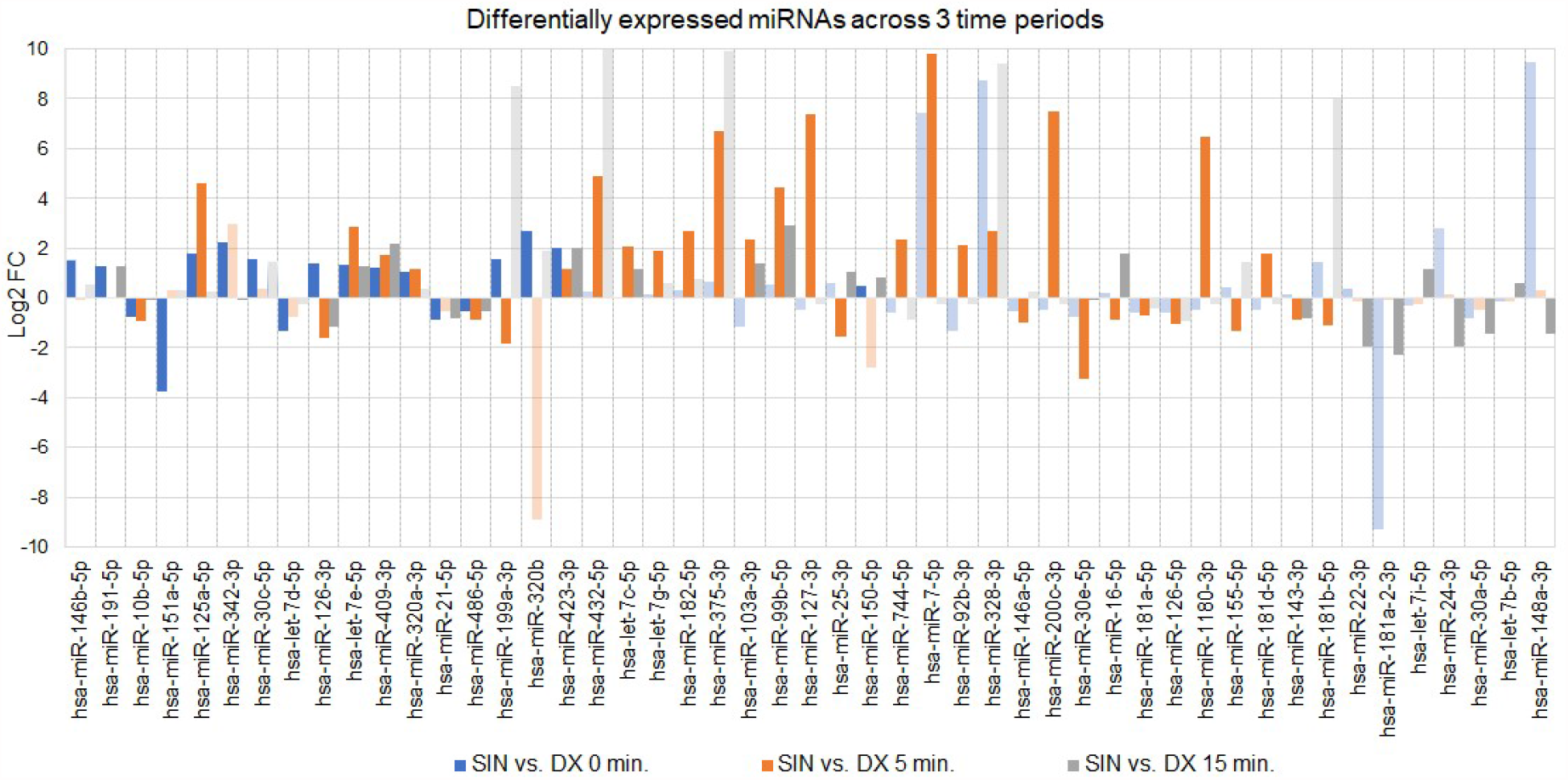
Differentially expressed miRNAs in BIPSS samples across all three time points. The bar chart represents Log2FCs of all differentially expressed miRNAs with Bonferroni P < 0.05. Transparent bars represent fold changes of adjacent time points (0, 5, 15) but are not statistically significant (Bonferroni P > 0.05). DX - dextral side, SIN - sinistral side.

In a time point of 0 minutes, the analysis revealed 17 DEMs. Of these 12 were upregulated while 5 were downregulated. In a time point of 5 minutes, the amount of differentially expressed miRNAs increased to 34 with 20 being upregulated and 14 being downregulated. As for the time point of 15 minutes, the amount of miRNAs decreased to 20 of which 11 were upregulated and 9 were downregulated (Figure 3). During the 5th minute of BIPSS, we detected two DEMs of interest: the aforementioned hsa-miR-375-3p (Log2FC: 6.7, 696.9 CPM vs 72625.89 CPM) and hsa-miR-7-5p which had the highest Log2FC of all DEMs (9.8, 21.8, CPM vs 20340.3 CPM). These two DEMs were also upregulated in plasma sample 24 hrs. after BIPSS with a Log2FC for hsa-miR-375-3p: 3.3 (20.4 CPM vs 208.8 CPM) and hsa-miR-7-5p: 1.1 (120.6 CPM vs 251.8 CPM). They were not differentially expressed in any of the other BIPSS time points. Further investigating these two miRNAs in PVB samples we detected < 106 read counts of hsa-miR-375-3p in all four samples which is notably lower than the 4581 read counts in the BIPSS-5-SIN sample. A similar result was observed with hsa-miR-7-5p as it had < 89 reads in all four PVB samples compared to the 1283 reads in BIPSS-5-SIN.

## 4. Discussion

Despite the vast advancements in the research of circulating miRNAs in cancer the studies of circulating miRNAs in PitNETs remain rare with only two published reports (Belaya et al., 2020; Németh et al., 2019). The reason for the scarce amount of studies could be that the hormone-secreting PitNETs with clinical manifestations are generally rare in the population (Daly and Beckers, 2020). Also, they are less proliferative than malignant tumors therefore once the PitNET secreted miRNAs are diluted within the blood their detection becomes difficult requiring highly sensitive methods. To our knowledge, this is the first study that has evaluated the circulating miRNAs in plasma samples that are acquired directly from BIPSS, a “gold standard” procedure in the diagnosis of ACTH secreting PitNETs. As a result, we were able to identify PitNET derived miRNAs that are secreted alongside with ACTH.

The first thing we observed was that BIPSS samples had about 3 times lower annotated (miRBase v22) read percentage. This could be due to the location of sample collection since the catheter is inserted close to the tumor itself. By mapping the reads to HG38 it can be observed that BIPSS samples had vastly more lncRNA reads than PVB samples (supplementary figure 8). It is known that lncRNAs can also be packed in EVs and secreted by the tumor and as a result they can modulate the tumor microenviroment to promote tumor growth (Zhou et al., 2020). Perhaps both PitNET and pituitary gland actively contributes to specific lncRNA secretion in plasma which would explain high amount of annotated lncRNA reads in BIPSS samples compared to PVB samples.

Another aspect regarding the annotated read percentage could be attributed to the miRNA extraction efficiency itself. During quality assessment by qPCR in PVB samples we observed lower Ct values of endogenous miRNAs as well as lower Ct values of UniSp100/UniSp101 spike-ins. It could be that plasma samples from BIPSS were spiked with inhibitors that might affect miRNA extraction itself because we extracted four BIPSS samples and four PVB samples at the same time (Supplementary Table 1). It is also important to note that BIPSS samples had only about 500 µl of plasma available per sample where PVB plasma was available at 1 ml per sample. In BIPSS the location of the sampling catheter is guided via transjugular approach to the

IPSs which are in close proximity to the pituitary gland. This allows to precisely assess the amount of secreted ACTH after the gland and PitNET is stimulated with CRH (Jarial et al., 2018). This in theory should increase the possibility to detect PitNET specific circulating miRNAs since their dilution is less than in blood sampled from PVB. More so a recent study published by Gümürdü A. showed for the first time that the stimulation of hormone release via exocytosis of large dense-core vesicles (LDCVs) is accompanied by the release of miRNAs. Their data concluded that in bovine chromaffin cells specific miRNAs (in their case miR-375) can be released upon stimulation alongside hormones and neurotransmitters (ribomones) (Gümürdü et al., 2017).

Our data strongly support the idea of ribomones as we observed a strong increase in plasma expression of two LDCV packed miRNAs hsa-miR-375-3p and hsa-miR-7-5p (Figure 4, Supplementary Table 11) within the sinistral side during 5 minutes of BIPSS. According to our clinical data, the tumor of the patient was located on the sinistral side and the highest amount of ACTH release was achieved during the 3^rd^ and 5^th^ minute of the procedure which was 24 times higher than in the dextral side (Table 1). Also, the amount of hsa-miR-375-3p and hsa-let-7-5p reads in PVB plasma before and after BIPSS was much lower than in the BIPSS-5-SIN sample further suggesting that they have potentially a PitNET origin and are specifically associated with the secretion of ACTH. This however raises the question of whether these two miRNAs have any impact on the clinical characteristics of the PitNET or are they purely associated with exocytosis of ACTH granules. hsa-miR-7-5p has been studied in non-small cell lung cancers, glioblastomas, and nasopharyngeal carcinomas with the majority of studies inclining towards hsa-miR-7-5p having tumor suppressing-characteristics (Xiao, 2019; Yin et al., 2019; Zhong et al., 2019). hsa-miR-375-3p has also been reported in the majority of studies as a tumor suppressive miRNA in cancers such as colorectal cancers, bladder cancers, and head and neck squamous cell carcinomas (Cen et al., 2018; Li et al., 2020; Xu et al., 2019). To further evaluate how these two miRNAs affect the phenotype of ACTH secreting PitNETs functional studies are needed.

A recently published study showed that plasma miRNAs miR-16-5p, miR-145-5p, and let-7g-5p are upregulated in CS patients with ACTH-secreting PitNETs versus CS patients with ectopic cause (Belaya et al., 2020). In our samples let-7g-5p was upregulated (Log2FC: 1.9) at the 5^th^ minute mark while miR-16-5p was found to be downregulated (Log2FC: −0.9). Interestingly, miR-16-5p was upregulated at the 15^th^ minute time point (Log2FC: 1.8) (Supplementary Table 11). In our study miR-145-5p was not differentially expressed in any of the time points. Upregulation of miR-16-5p and let-7g-5p within the sinistral side at 15 minutes after stimulation by CRH could indeed indicate that these miRNAs might be of PitNET origin, however functional studies are required validate this claim.

As for the top ten most expressed miRNAs in BIPSS and PVB samples we compared our dataset with the dataset that contained plasma miRNAs NGS data from FSH/LH PitNET patients (Németh et al., 2019). With this, we further deduced whether the top ten most miRNAs expressed miRNAs are related to the tumor or they are constitutively expressed in plasma. We found that hsa-miR-486-5p was the 2^nd^ most expressed miRNA and that six of the remaining nine most expressed miRNAs in the BIPSS plasma sample group could also be found amongst the 20 most expressed miRNAs in FSH/LH dataset (Supplementary Table 13, Figure 1). The exceptions are: hsa-miR-10b-5p, hsa-miR-375-3p, hsa-miR10a-5p. As for the 24 hrs. before and after BIPSS samples we discovered that 8 out of 10 most expressed miRNAs in our data were also amongst the 20 most expressed miRNAs in FSH/LH dataset (exceptions: hsa-miR-10b-5p, hsa-miR-181a-5p). The same result was also obtained for the samples before and after surgery.

In the literature, there is evidence of hsa-miR-486-5p having both tumor-suppressive and tumor-promoting effects in tissue studies (Tian et al., 2019a; Yang et al., 2017). However, there is also evidence for hsa-miR-486-5p having an erythropoietic origin and should be interpreted with caution in plasma miRNA studies (Juzenas et al., 2020). Similar results were also observed with hsa-let-7(a,b,f)-5p, hsa-miR-92a-3p, hsa-miR-30d-5p, hsa-miR-26a-5p as they were also found amongst the top 20 most expressed miRNAs in FSH/LH dataset suggesting they are constitutively expressed in plasma and most likely do not represent the secreted miRNAs by PitNET. However, there is also strong evidence for miRNAs of the let-7 family having an involvement in tumorigenesis by acting primarily as tumor suppressors (Lee et al., 2016). In our dataset, only hsa-miR-7b-5p was differentially expressed with a negative fold change (Figure 4). The same can be said about hsa-miR-486-5p as it’s been shown to suppress tumor growth in non-small cell lung cancer by targeting PIK3A1 (Tian et al., 2019b). Indeed, in our samples, we observed decreased expression of hsa-miR-486-5p within the sinistral side in all three time points (Figure 4).

## 5. Conclusions

This study has provided the first insight into plasma miRNome from samples that are acquired from the BIPSS procedure allowing us to investigate PitNET secreted miRNAs in close proximity to the tumor itself. We were able to distinguish BIPSS sample miRNA composition from PVB fraction. Furthermore, the results indicate that hsa-miR-7-5p and hsa-miR-375-3p are directly released upon the secretion of ACTH suggesting that these two miRNAs can serve as potential biomarkers and further functional characterization of these miRNA are needed to estimate the role in PitNET pathogenesis.

## Data Availability

Data supporting the findings that are reported within the manuscript are available in supplementary files. Sequencing data are available in FASTQ file format at Gene Expression Omnibus public repository with accession number: GSE163957.
Any additional data is available from corresponding authors upon request.

https://www.ncbi.nlm.nih.gov/geo/query/acc.cgi?acc=GSE163957

## Abbreviations

miRNA: micro RNA
PitNET: pituitary neuroendocrine tumor
BIPSS: bilateral inferior petrosal sinus sampling
IPS: inferior petrosal sinus
PVB: peripheral venous blood
CRH: corticotropin-releasing hormone
hCRH: human corticotropin-releasing hormone
ACTH: adrenocorticotropic hormone
CD: Cushing’s disease
MRI: magnetic resonance imaging
FSH/LH: Follicle-stimulating hormone / luteinizing hormone
EV: extracellular vesicle
NGS: next-generation sequencing
EDTA: ethylenediaminetetraacetic acid
qPCR: quantitative polymerase chain reaction
Log2FC: Log2 of fold change

## Data Availability

Data supporting the findings that are reported within the manuscript are available in supplementary files. Sequencing data are available in FASTQ file format at Gene Expression Omnibus public repository with accession number: GSE163957.

Link: https://www.ncbi.nlm.nih.gov/geo/query/acc.cgi?acc=GSE163957.

Any additional data is available from corresponding authors upon request.

## Ethics Statement

The use of patient’s samples and data in this study was approved by the Central Medical Ethics Committee of Latvia (protocol: 22.03.07/A7 and 2/18-02-21). The patients/participants provided their written informed consent to participate in this study.

## Author Contributions

HN performed RNA extraction, library preparation, NGS data analysis. Manuscript was prepared by HN, RP, VR. Patients clinical data was provided by IK and MR. All the figures and tables within manuscript and supplementary were prepared by HN and reviewed by RP and VR. Patient management and recruitment was done by IK, IB, MR, LS, JS, JS. All authors have read the final manuscript and approved it.

## Acknowledgments

This research was supported by the European Regional Development Fund within the project “RNA molecular determinants in development of pituitary adenoma” (1.1.1.1/18/A/089). The authors acknowledge the Latvian Biomedical Research and Study Centre and the Genome Database of the Latvian Population for providing infrastructure, biological material and data.

