## supplementary figure for "Micro-RNAs miR-375-3p and miR-7-5p are released alongside ACTH from corticotroph pituitary neuroendocrine tumor"

Supplementary Material

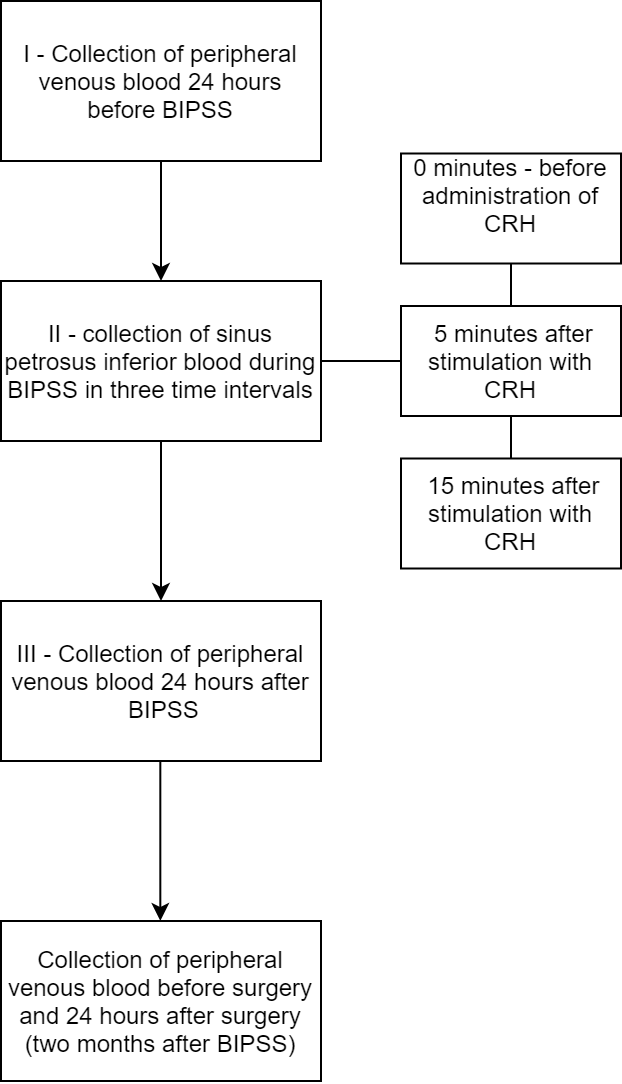

**Supplementary Figure 1.** Collection of plasma samples used in miRNA extraction and NGS

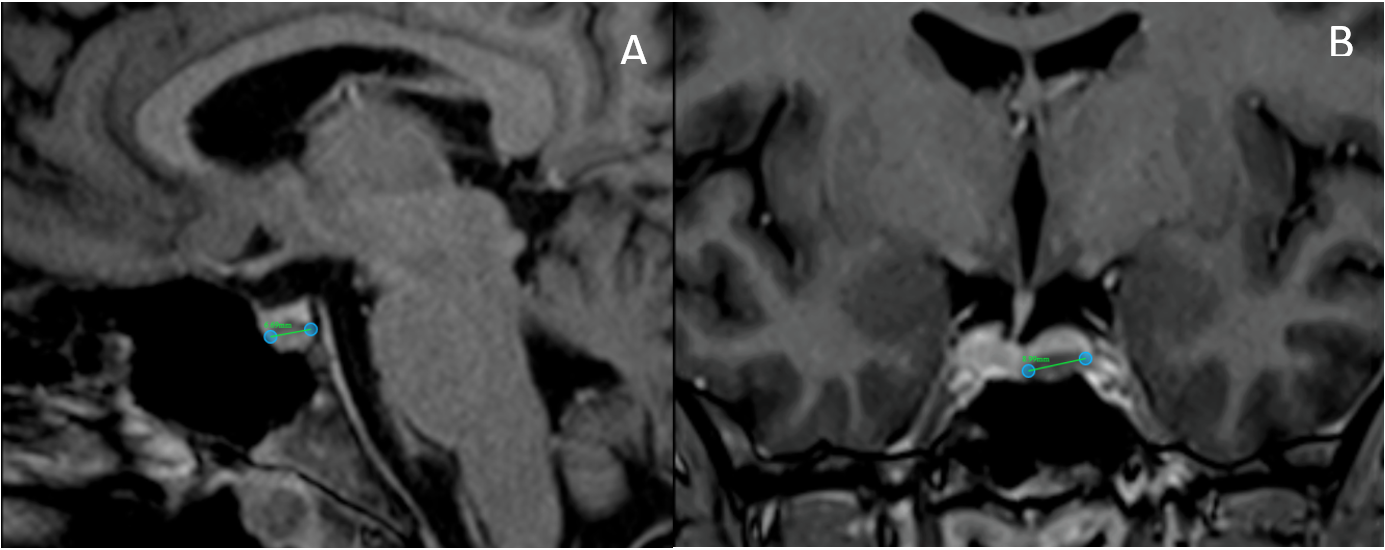

**Supplementary Figure 2.** T1-weighted MRI of pituitary, sagittal (A) and coronal (B) planes.

UniSp 100 / UniSp101

cT

|  | UniSp100 | UniSp101 |
| --- | --- | --- |
| BIPSS-0-DX | 30.3689 | 27.1551 |
| BIPSS-0-SIN | 30.3475 | 27.3546 |
| BIPSS-5-DX | 31.1246 | 27.9405 |
| BIPSS-5-SIN | 32.0792 | 28.6926 |
| BIPSS-15-DX | 31.0161 | 27.8098 |
| BIPSS-15-SIN | 29.3707 | 26.0688 |
| PVB-24 hrs. Before BIPSS | 28.5368 | 24.8178 |
| PVB-24 hrs. After BIPSS | 28.5637 | 25.0476 |
| PVB-before surgery | 28.7722 | 25.1807 |
| PVB-24 hrs. after surgery | 28.6327 | 24.8349 |

dcT UniSp100/UniSp101

|  | dcT (UniSp100 - UniSp101) |
| --- | --- |
| BIPSS-0-DX | 3.21373 |
| BIPSS-0-SIN | 2.99289 |
| BIPSS-5-DX | 3.1841 |
| BIPSS-5-SIN | 3.38657 |
| BIPSS-15-DX | 3.20631 |
| BIPSS-15-SIN | 3.30188 |
| PVB-24 hrs. Before BIPSS | 3.71899 |
| PVB-24 hrs. After BIPSS | 3.51615 |
| PVB-before surgery | 3.59147 |
| PVB-24 hrs. after surgery | 3.79781 |

**Supplementary Figure 3.** Ct Values and ΔCt values of UniSp100/UniSp101 spike-in qPCR. According to the user manual of used kit (QIAseq miRNA Library QC qPCR Assay Kit, Qiagen, Germany) the Ct values of UniSp100/UniSp100 should not vary by more than 3 between samples. The ΔCt values (UniSp100 – UniSp101) should be between 5 and 7.

UniSp6

cT

|  | 18,48 18,32 19,94 19,97 18,40 18,22 18,09 18,21 19,84 18,21 |
| --- | --- |
| BIPSS-0-DX | 18.4805 |
| BIPSS-0-SIN | 18.3158 |
| BIPSS-5-DX | 19.9404 |
| BIPSS-5-SIN | 19.9665 |
| BIPSS-15-DX | 18.4003 |
| BIPSS-15-SIN | 18.2187 |
| PVB-24 hrs. Before BIPSS | 18.0853 |
| PVB-24 hrs. After BIPSS | 18.2113 |
| PVB-before surgery | 19.8416 |
| PVB-24 hrs. after surgery | 18.2149 |

**Supplementary Figure 4.** Ct values of UniSp6 to evaluate synthesis efficiency of cDNA used in quality control qPCR. According to the user manual of used kit (QIAseq miRNA Library QC qPCR Assay Kit, Qiagen, Germany) the Ct values of UniSp6 should be < 20.

Haemolsysis assessment

|  | dcT (miR23a-3p - miR-451a) |
| --- | --- |
| BIPSS-0-DX | 6.05929 |
| BIPSS-0-SIN | 5.14975 |
| BIPSS-5-DX | 1.79016 |
| BIPSS-5-SIN | 3.33449 |
| BIPSS-15-DX | 3.16219 |
| BIPSS-15-SIN | 3.23586 |
| PVB-24 hrs. Before BIPSS | 3.83304 |
| PVB-24 hrs. After BIPSS | 3.74849 |
| PVB-before surgery | 2.72564 |
| PVB-24 hrs. after surgery | 2.88814 |

**Supplementary Figure 5.** Hemolysis assessment (ΔCt values of miR-23a-3p – miR-451a). According to the user manual the of used kit (QIAseq miRNA Library QC qPCR Assay Kit, Qiagen, Germany) the ΔCt values of <5 represent no hemolysis risk, 5 – 7 represent low hemolysis risk, >7 represent hemolyzed sample.

Endogenous miRNA controls assessment

cT

|  | miR-103a-3p | miR-191-5p | miR-30c-5p |
| --- | --- | --- | --- |
| BIPSS-0-DX | 34.8781 | 31.5528 | 32.2608 |
| BIPSS-0-SIN | 31.2867 | 30.4422 | 31.6147 |
| BIPSS-5-DX | 33.6399 | 29.645 | 32.718 |
| BIPSS-5-SIN | 36.2047 | 31.3105 | 33.5393 |
| BIPSS-15-DX | 34.9906 | 30.4282 | 32.167 |
| BIPSS-15-SIN | 28.5029 | 28.0922 | 29.5918 |
| PVB-24 hrs. Before BIPSS | 25.5195 | 25.9823 | 27.0994 |
| PVB-24 hrs. After BIPSS | 25.7313 | 26.0525 | 26.293 |
| PVB-before surgery | 24.5419 | 24.8918 | 26.293 |
| PVB-24 hrs. after surgery | 24.5577 | 24.8992 | 26.2912 |

**Supplementary Figure 6.** Endogenous miRNA controls assessment. According to the user manual of the used kit (QIAseq miRNA Library QC qPCR Assay Kit, Qiagen, Germany) if the extraction was successful these three miRNAs should be detected in plasma at any Ct value.

| **Before trimming** | **After trimming** |
| --- | --- |
| 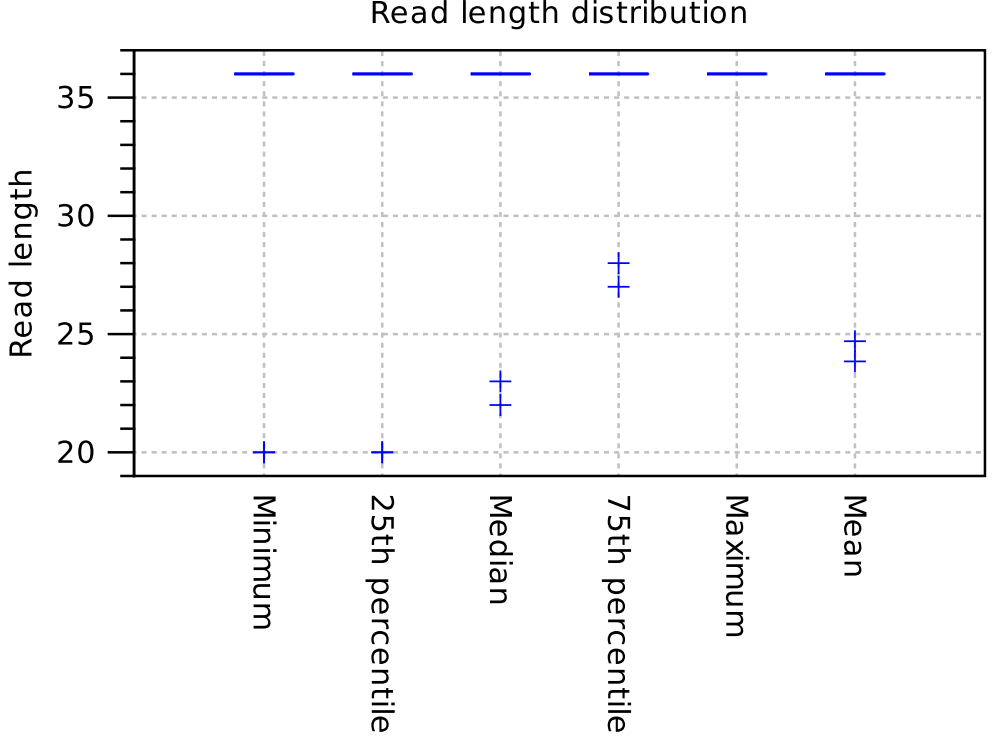 | 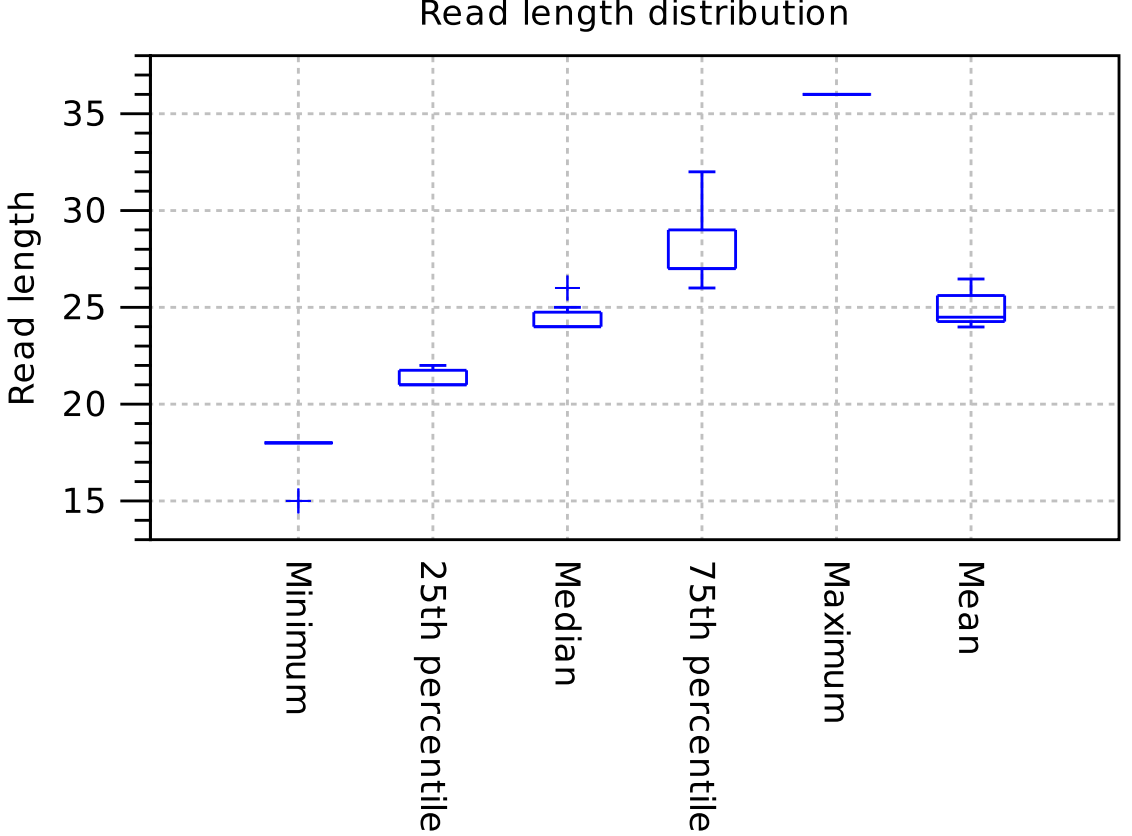 |
| 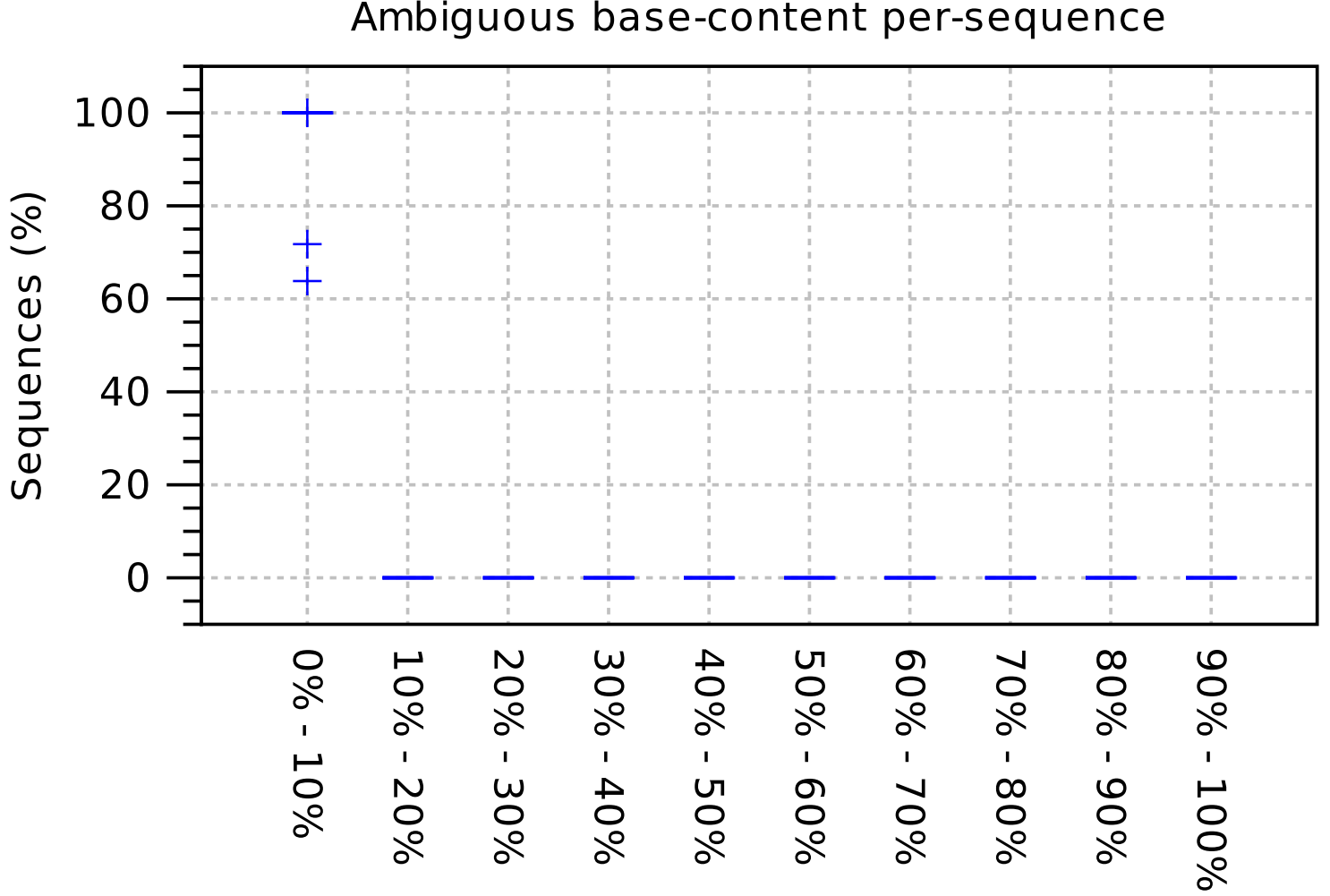 | All reads with ambigous bases were discared.  No data available. |
| 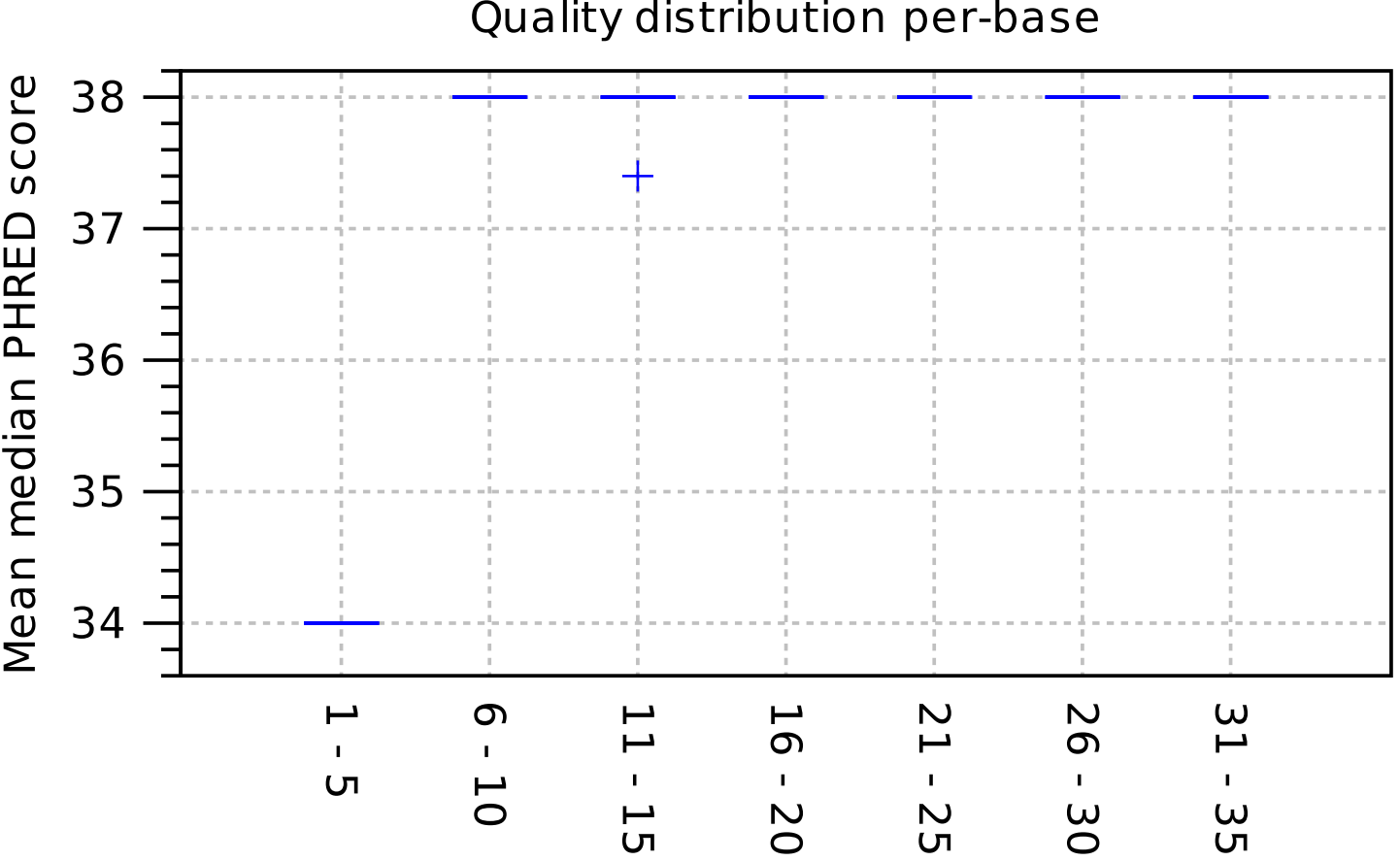 | 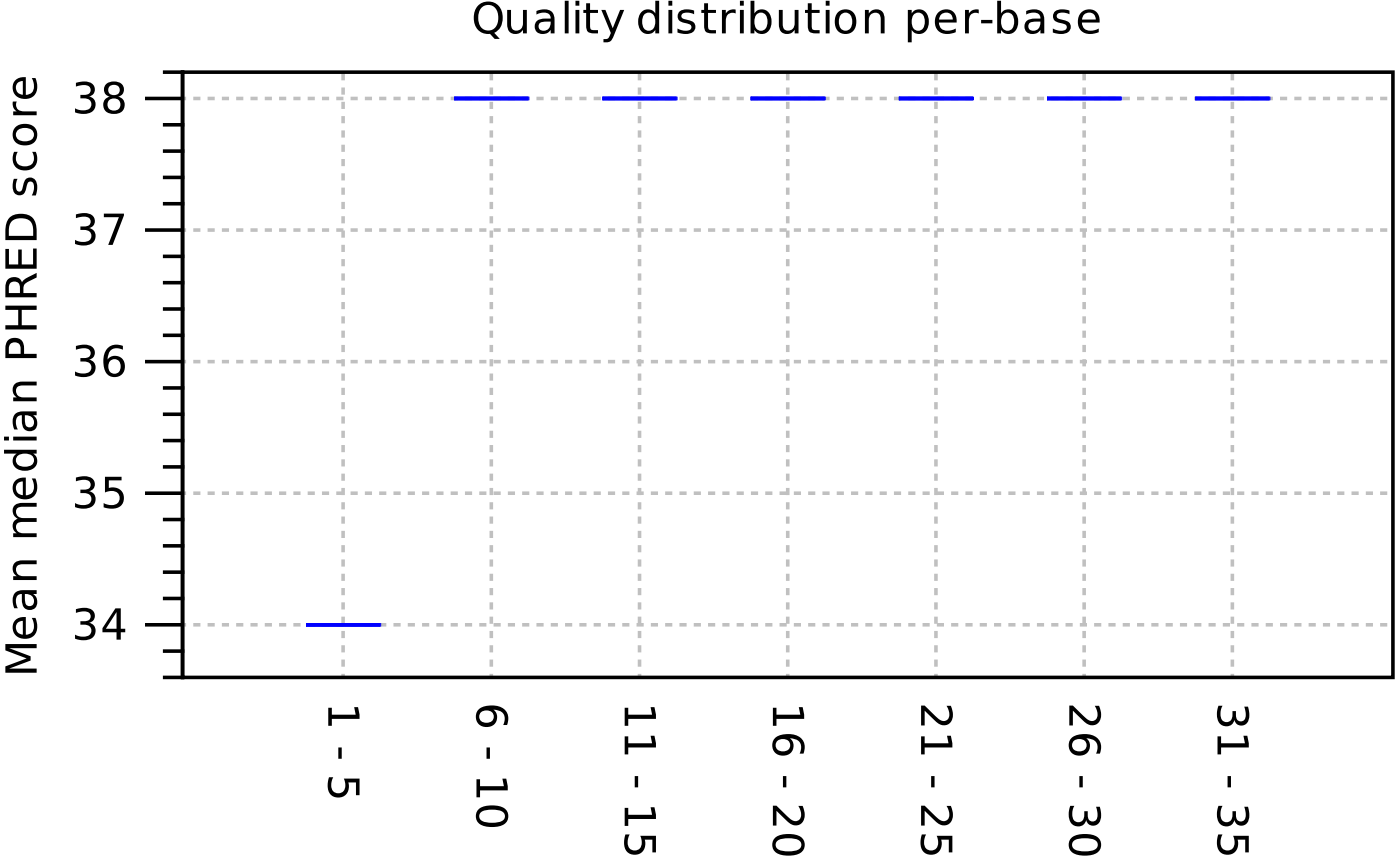 |
| 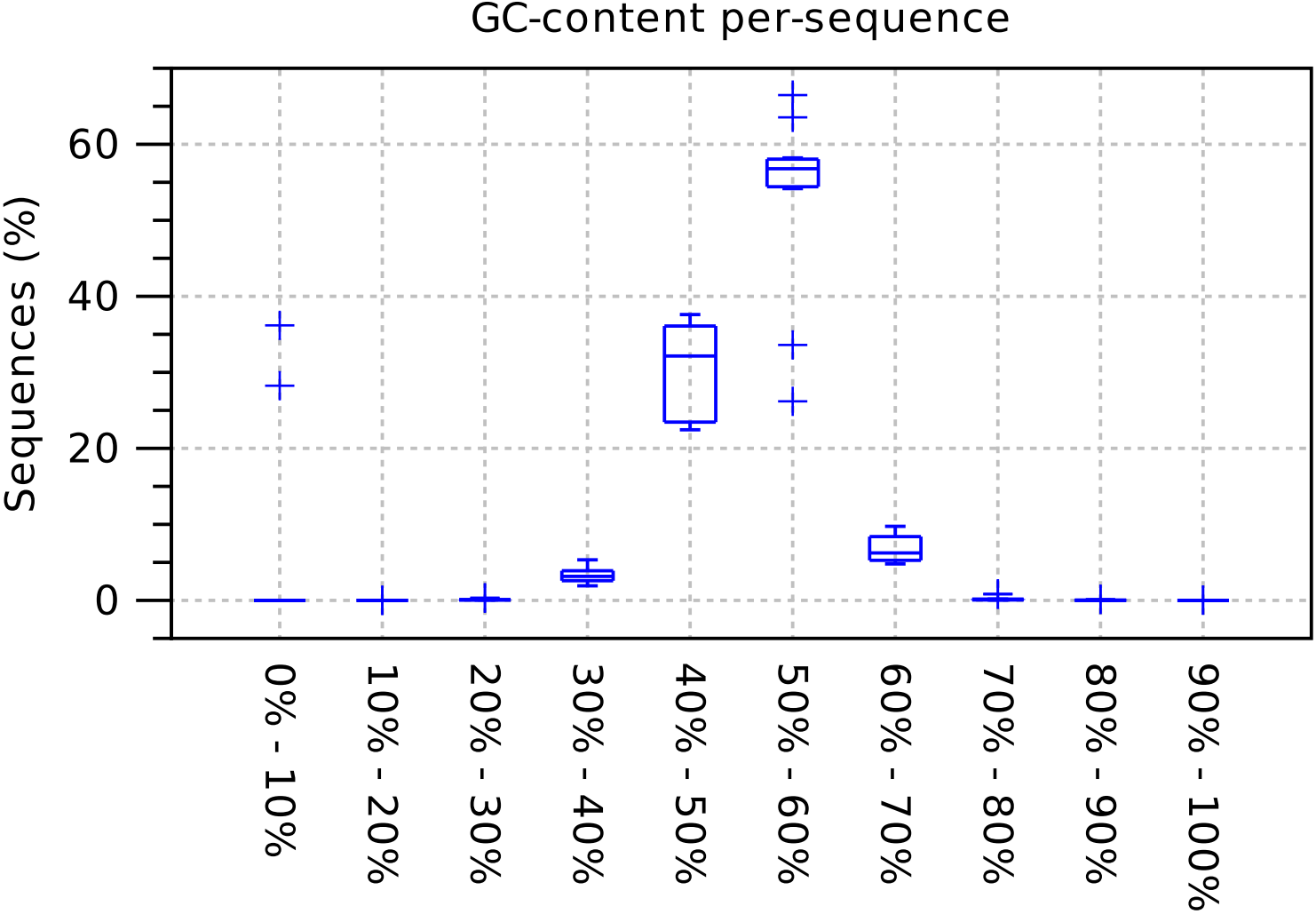 | 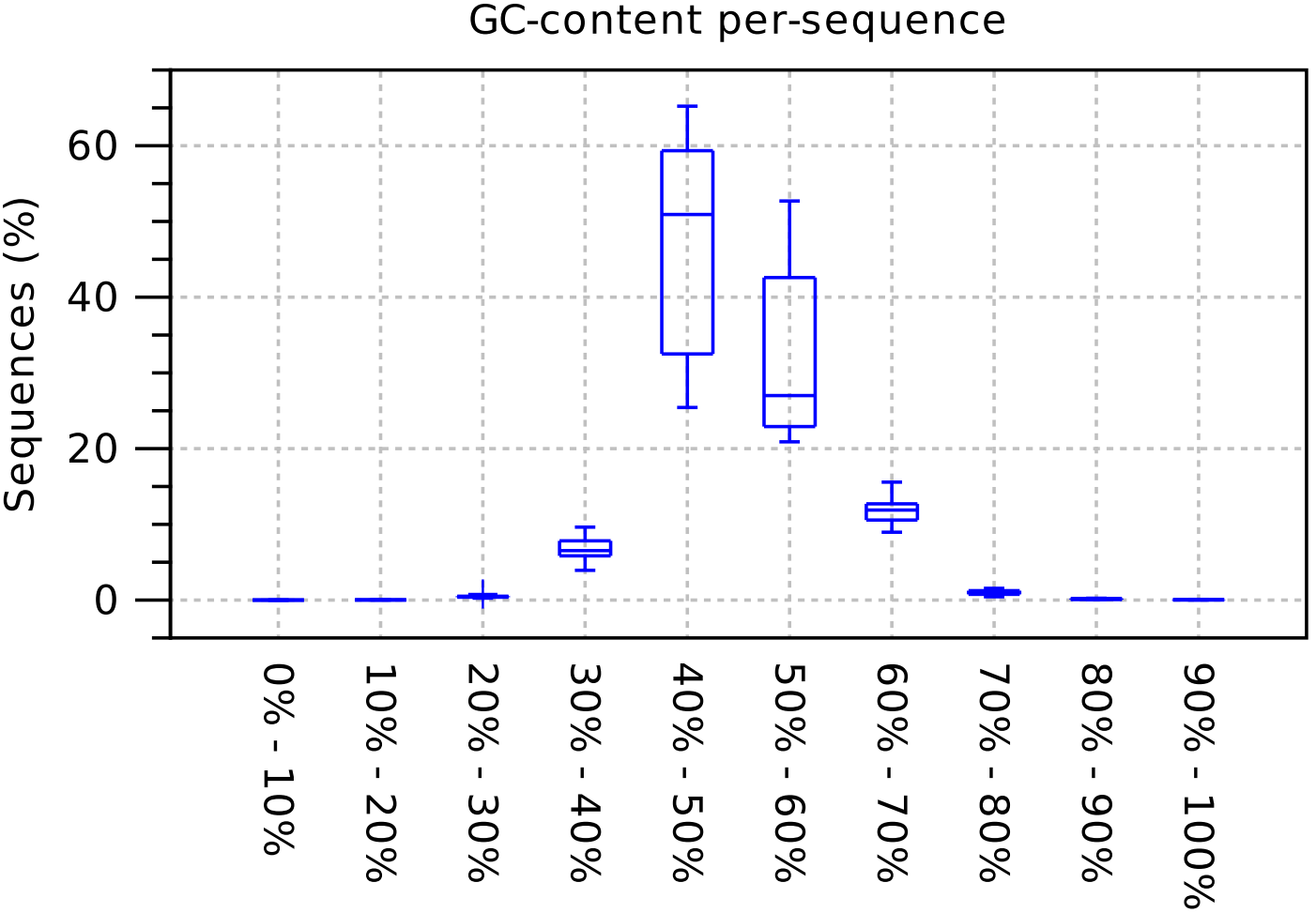 |

**Supplementary Figure 7 –** summary of sequence quality statistics before and after trimming.

**Read length distribution:** The box plot is based on 10 samples.

**Ambiguous base-content:** Summarizes the distribution of N-contents. The N-content of a sequence is calculated as the number of ambiguous bases compared to all bases. x: p% - r%: percentage range y: the number of sequences featuring particular N-percentages normalized to the total number of sequences The box plot is based on 10 samples.

**Quality distribution:** Summarizes the distribution of average sequence quality scores. The quality of a sequence is calculated as the arithmetic mean of its base qualities. x - y: PHRED range y: the number of sequences observed at that qual. score normalized to the total number of sequences The box plot is based on 10 samples.

**GC-content:** Summarizes the distribution of GC-contents. The GC-content of a sequence is calculated as the number of GC-bases compared to all bases (including ambiguous bases). x: p% - r%: percentage range y: the number of sequences featuring the particular GC-percentage range normalized to the total number of sequences The box plot is based on 10 samples.

Read mapping to HG38

|  | miRNA | lncRNA | protein coding | nonsense_mediated_decay | misc_RNA | retained_intron | processed_transcript | sno_RNA | rRNA | snRNA | Mt_tRNA | sca_RNA | Mt_rRNA | sRNA | non_stop_decay | polymorphic_pseudogene | unknown | ribozyme |
| --- | --- | --- | --- | --- | --- | --- | --- | --- | --- | --- | --- | --- | --- | --- | --- | --- | --- | --- |
| BIPSS-0-DX | 28.79% | 24.32% | 16.93% | 13.19% | 6.86% | 4.1% | 2.82% | 0.26% | 0.11% | 0.1% | 0.07% | 0.04% | 0.01% | 0.01% | 0% | 0% | 2.39% | 0% |
| BIPSS-0-SIN | 21.72% | 28.58% | 17.5% | 13.92% | 8.67% | 4.01% | 2.99% | 0.22% | 0.05% | 0.15% | 0.12% | 0.02% | 0.01% | 0% | 0% | 0% | 2.04% | 0% |
| BIPSS-5-DX | 21.53% | 21.12% | 14% | 20.04% | 13.08% | 3.64% | 2.91% | 0.28% | 0.09% | 0.13% | 0.11% | 0.02% | 0.03% | 0% | 0% | 0% | 3.02% | 0% |
| BIPSS-5-SIN | 26.63% | 18.16% | 12.32% | 19.69% | 13.13% | 2.9% | 2.5% | 1.39% | 0.2% | 0.2% | 0.48% | 0.03% | 0.14% | 0% | 0% | 0% | 2.23% | 0% |
| BIPSS-15-DX | 20.5% | 27.57% | 15.95% | 17.15% | 8.76% | 4.72% | 3.04% | 0.23% | 0.04% | 0.12% | 0.09% | 0.02% | 0% | 0% | 0% | 0% | 1.81% | 0% |
| BIPSS-15-SIN | 19.53% | 31.53% | 20.58% | 6.9% | 10.27% | 4.67% | 3% | 0.37% | 0.12% | 0.06% | 0.12% | 0.01% | 0.02% | 0% | 0.01% | 0.01% | 2.8% | 0% |
| PVB-24 hrs. Before BIPSS | 29.07% | 2.98% | 3.49% | 1.44% | 57.89% | 0.82% | 0.61% | 0.24% | 0.09% | 0.01% | 0.3% | 0.02% | 0.1% | 0% | 0% | 0% | 2.94% | 0% |
| PVB-24 hrs. After BIPSS | 29.76% | 7.1% | 8.25% | 3.63% | 40.64% | 2.08% | 1.44% | 0.28% | 0.21% | 0.02% | 0.2% | 0.07% | 0.07% | 0% | 0% | 0% | 6.25% | 0% |
| PVB-before surgery | 31.05% | 6.83% | 7.68% | 6.56% | 35.42% | 1.87% | 1.42% | 0.65% | 0.17% | 0.03% | 0.37% | 0.02% | 0.07% | 0% | 0.01% | 0% | 7.84% | 0.01% |
| PVB-24 hrs. after surgery | 24.36% | 5.72% | 5.51% | 3.51% | 50.78% | 1.3% | 0.99% | 0.63% | 0.1% | 0.02% | 0.4% | 0.02% | 0.1% | 0% | 0.01% | 0% | 6.55% | 0% |

**Supplementary Figure 8.** Read mapping to human genome (hg38) with CLC Genomics Workbench (V20.0.4.). The graph represents distribution of biotypes in each sample.
